# A Model to Predict In-hospital Mortality in Elderly Patients with Community-acquired Pneumonia: a Retrospective Study

**DOI:** 10.1101/2022.04.18.22273956

**Authors:** Yanting Hao, Hua Zhang, Yingying Yan, Yun Zhu, Fuchun Zhang

## Abstract

**Objectives:** To develop and validate a predictive model for evaluating in-hospital mortality risk in elderly patients with community-acquired pneumonia.

**Setting:** Two tertiary care hospitals with 2000 beds and 1000 beds respectively in Beijing, China.

**Participants:** Elderly patients (age ≥ 65 years) with community-acquired pneumonia admitted to the Department of Internal Medicine of the two hospitals from January 2010 to December 2019 or from January 2019 to December 2019 respectively.

**Design:** It was a retrospective study. After dividing the data set into training and validation sets, we created a mortality model that included patient demographics, hospitalization time, hospital outcome, and various clinical conditions associated with hospitalization. We then applied the model to the validation set.

**Main outcome measures:** In-hospital mortality.

**Results:** The training cohort included 2,466 patients admitted to the one hospital, while the validation cohort included 424 patients at the other hospital. The overall in-hospital mortality rate was 15.6%. In the multivariable model, age, respiratory failure, heart failure, and malignant tumors were associated with mortality. The model had excellent discrimination with AUC values of 0.877 and 0.930 in the training and validation cohorts, respectively.

**Conclusions:** The predictive model to evaluate in-hospital mortality in elderly patients with community-acquired pneumonia, which was established based on administrative data, provides a simple tool for physicians to assess the prognosis of elderly patients with community-acquired pneumonia in Beijing.

**Strengths and limitations of this study:**

1. The clinical data collected were obtained from a large retrospective cohort over a 10-year period, resulting in good reliability.
2. The model based on administrative data can help healthcare workers determine the prognosis and outcomes of elderly patients of CAP, especially in resource limited regions in China.
3. Our main outcome was in-hospital mortality, not 30-day mortality or longer.
4. All included cases were collected from inpatient, outpatient and emergency patients were not included.
5. The model was only verified in two hospitals in Beijing and further verification should be conducted in other areas and different levels of hospitals.

## INTRODUCTION

Community-acquired pneumonia (CAP) is an important threat to the health of elderly adults.^1^ Like other respiratory infections, people at the extremes of age are at greatest risk for CAP and have the worst outcomes. In developed countries, almost half of all hospitalizations for CAP occur in patients over 65, and pneumonia is a leading cause of death among this age group.^2^ In China, the in-hospital mortality rate of CAP in 95-year-olds is as high as 36.1%.^3^ This high mortality indicates the importance of identifying patients at high risk of death so that clinical intervention can be provided early on. Therefore, it is crucial to predict the mortality of elderly patients with CAP.

The pneumonia severity index (PSI) is a widely accepted and validated severity scoring system that assesses the risk of mortality for pneumonia patients in a two-step algorithm.^4^ Unfortunately, the PSI is a complex metric that involves 20 variables, limiting its dissemination and implementation in everyday practice. In addition, the PSI is best for assessing patients with low mortality risk who may be suitable for home management rather than elderly patients with severe CAP. The CURB-65 score, which is based on Confusion, Urea, Respiratory rate, and Blood pressure, has been proposed as a simpler alternative to PSI.^5^ However, this score also has limitations. CURB-65 stratifies patients into only two groups (severe and non-severe); it does not identify patients at low risk of mortality who might be suitable for early hospital discharge or home management. Therefore, both PSI and CURB-65 have limitations.

Elderly patients with CAP have many comorbidities, and the cases can be critical and complicated. The development of a tool to predict the outcomes of CAP earlier would allow the management strategy to be adjusted sooner, which would help prevent deterioration and improve the survival rate. In clinical work, data including patients’ clinical diagnoses and comorbidities can be easily obtained. A simple and quick tool that uses these accessible data to predict the prognosis of elderly CAP patients during hospitalization would be very helpful.

The aim of this study was to develop an assessment tool that can be used to stratify elderly patients hospitalized with CAP into groups with different mortality risk based on administrative data.

## METHODS

### Study Population

This was a retrospective study. The patients admitted to the one hospital (A) from January 01, 2010 to December 31, 2019 and the other hospital (B) from January 01, 2019 to December 31, 2019 were included. The inclusion criteria were as follows: (1) age ≥ 65 years; (2) main diagnosis (i.e., the cause of admission) of CAP; and (3) inpatients admitted to the Department of Internal Medicine of the two hospitals. The exclusion criteria were SARS and suspected patients and patients with active pulmonary tuberculosis, HIV positive, organ transplantation, obstructive pneumonia, lung abscess, radiation pneumonia, or *Pneumocystis carinii* pneumonia.

### Data Collection

We collected social demographic data including age and gender along with clinical data including hospitalization time, hospital outcome, and concomitant diseases (e.g., thrombotic diseases, stress ulcers, catheter related infections, delirium, adverse drug reactions, malnutrition, respiratory failure, heart failure, chronic lung diseases, malignant tumors, and acute myocardial infarction) during hospitalization. The prognosis was recorded as either survival (when the patient was discharged from the hospital) or death.

### Training and Validation Cohorts

The patients from the hospital A were assigned to the training cohort, while the population from the hospital B served as the validation cohort. The training cohort was used to develop the prediction model, and external verification was carried out using the validation cohort.

### Statistical Analysis

All data were analyzed using SPSS 20.0 (IBM, New York, USA). Normally distributed continuous variables were expressed as mean ± standard deviation, and independent sample t-test was used for comparison between groups. Variables that did not conform to the normal distribution were expressed as the median (25% quantile, 75% quantile), and Wilcox test was used for comparison between groups. The classified variables were statistically described by frequency (percentage), and the differences between groups were compared by χ2 test or Fisher’s exact test. The logistic regression method was established with hospital death as the dependent variable and potential influencing factors with *P* < 0.2 in the single-factor analysis as independent variables; regression was then used to screen the variables. A two-sided *P*-value less than 0.05 was considered to indicate a statistically significant difference.

According to the multi-factor analysis results combined with expert opinions, the predictive factors with clinical significance were selected to establish a predictive model based on logistic regression, and a line chart was established based on the regression results. The training cohort was used for internal verification, while the validation cohort is used for external verification. The areas under the ROC curve (AUC), correction curve, and clinical decision curve were calculated to determine the predictive ability of the model. Excellent predictive ability was indicated by AUC > 0.85, while AUC in the range of 0.7–0.85 indicated acceptable predictive ability.

### Patient and public involvement

Patients and the public were not involved in the study design, recruitment or execution, and the participants were not to be involved in disseminating the results.

## RESULTS

### Patient Features

The training and validation sets included 2,466 and 424 patients, respectively. The characteristics of these patients are listed in Table 1. In the training set, the majority of patients were male (64.52%), the average age was 80 years (range of 74–85 years), the average hospital stay was 13 days (range of 9–21 days), and hospital death occurred in 17.36% of patients. In the validation cohort, 55.19% of the patients were male, the average age was 83 years (78–88 years), the average hospital stay was 12 days (8–16 days), and the in-hospital mortality rate was 5.66%. The concomitant clinical conditions can be found in Table 1.

**Table 1.** Baseline characteristics of the training set and validation set

|  | Training set (*n* = 2,466) | Validation set (*n* = 424) |
| --- | --- | --- |
| Gender |  |  |
| Male | 1591 (64.52%) | 234 (55.19%) |
| Female | 875 (35.48%) | 190 (44.81%) |
| Age | 80 (74–85) | 83 (78–88) |
| Day | 13 (9–21) | 12 (8–16) |
| Thromboembolic disease | 125 (5.07%) | 41 (9.67%) |
| Pressure ulcer | 79 (3.20%) | 17 (4.01%) |
| Catheter related urinary tract infection | 45 (1.82%) | 12 (2.83%) |
| Delirium | 14 (0.57%) | 2 (0.47%) |
| Side effects from medication | 31 (1.26%) | 10 (2.36%) |
| Malnutrition | 363 (14.72%) | 133 (31.37%) |
| Respiratory failure | 608 (24.66%) | 81 (19.10%) |
| Heart failure | 531 (21.53%) | 122 (28.77%) |
| Chronic pulmonary disease | 602 (24.41%) | 156 (36.72%) |
| Cancer | 381 (15.45%) | 52 (12.26%) |
| Acute myocardial infarction | 54 (2.19%) | 3 (0.71%) |
| In-hospital mortality | 428 (17.36%) | 24 (5.66%) |

### Single-factor analysis

Based on univariate analysis in the training set, compared to surviving patients, the patients who died were significantly higher in age (*P* < 0.001), had a significantly longer hospital stay (*P* < 0.001), and incurred a significantly higher cost of hospitalization (*P* < 0.001). The incidences of the following diseases were also significantly different between dying and surviving patients (Table 2): thrombotic disease (*P* < 0.024), stress ulcer (*P* < 0.001), catheter-related infection (*P* < 0.014), malnutrition (*P* < 0.001), respiratory failure (*P* < 0.001), heart failure (*P* < 0.001), tumors (*P* < 0.001), and acute myocardial infarction (*P* < 0.001).

**Table 2.**
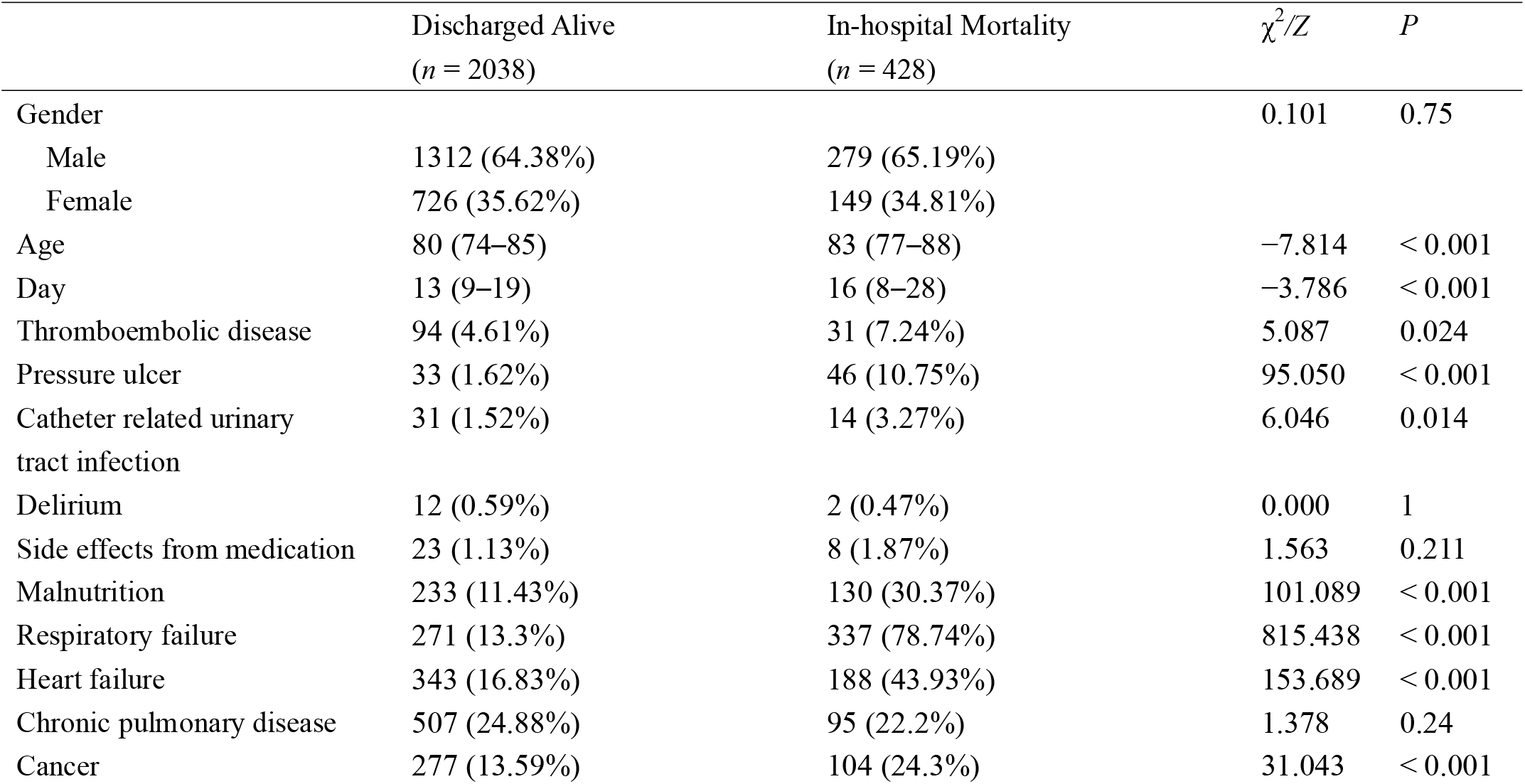
Comparison of two groups (discharged alive and in-hospital mortality) in the training set

| | Discharged Alive<br>($n = 2038$) | In-hospital Mortality<br>($n = 428$) | $\chi^2/Z$ | $P$ |
| --- | --- | --- | --- | --- |
| Gender |  |  | 0.101 | 0.75 |
| Male | 1312 (64.38%) | 279 (65.19%) |  |  |
| Female | 726 (35.62%) | 149 (34.81%) |  |  |
| Age | 80 (74–85) | 83 (77–88) | −7.814 | < 0.001 |
| Day | 13 (9–19) | 16 (8–28) | −3.786 | < 0.001 |
| Thromboembolic disease | 94 (4.61%) | 31 (7.24%) | 5.087 | 0.024 |
| Pressure ulcer | 33 (1.62%) | 46 (10.75%) | 95.050 | < 0.001 |
| Catheter related urinary tract infection | 31 (1.52%) | 14 (3.27%) | 6.046 | 0.014 |
| Delirium | 12 (0.59%) | 2 (0.47%) | 0.000 | 1 |
| Side effects from medication | 23 (1.13%) | 8 (1.87%) | 1.563 | 0.211 |
| Malnutrition | 233 (11.43%) | 130 (30.37%) | 101.089 | < 0.001 |
| Respiratory failure | 271 (13.3%) | 337 (78.74%) | 815.438 | < 0.001 |
| Heart failure | 343 (16.83%) | 188 (43.93%) | 153.689 | < 0.001 |
| Chronic pulmonary disease | 507 (24.88%) | 95 (22.2%) | 1.378 | 0.24 |
| Cancer | 277 (13.59%) | 104 (24.3%) | 31.043 | < 0.001 |
| Acute myocardial infarction | 34 (1.67%) | 20 (4.67%) | 14.909 | < 0.001 |

### Multifactor analysis

Taking hospital death as a dependent variable and the potential influencing factors with *P* < 0.2 in the single-factor analysis as independent variables, we established a logistic regression model and screened the variables via regression. Multivariate logistic regression analysis showed that age (OR = 1.03, 95% CI 1.0–1.1, *P* = 0.001), respiratory failure (OR = 20.3, 95% CI 15.5–26.7, *P* < 0.001), heart failure (OR = 2.2, 95% CI 1.7–2.9, *P* < 0.001), and tumors (OR = 2.3, 95% CI 1.6–3.1, *P* < 0.001) were independent risk factors of hospital death in elderly patients with CAP (Table 3).

**Table 3.**
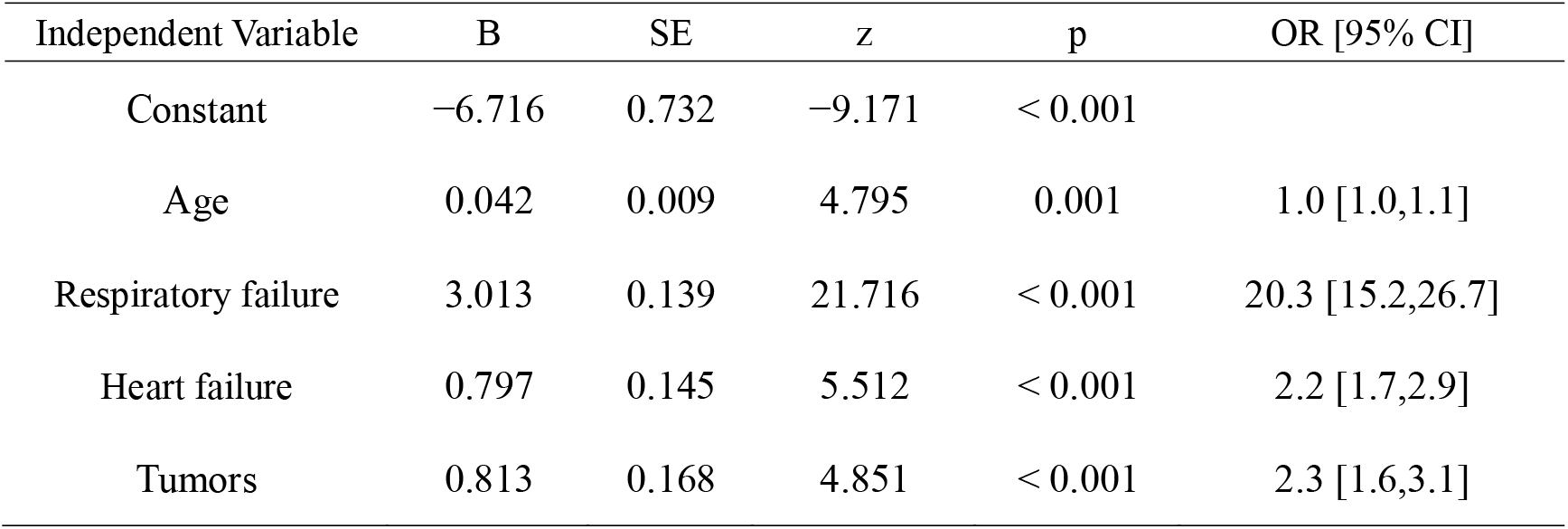
Multivariate logistic regression analysis of the training set

### Development of the Predictive Model

Based on the results of the above multifactor analysis, the statistically significant factors (age, respiratory failure, heart failure, and tumors) were taken as predictive factors to establish the prediction model. A nomogram is created as Figure 1. To predict a patient’s mortality rate, the point on each line is obtained according to the patient’s characteristics, and the patient’s total score is calculated, According to the total score, the corresponding mortality rate can be queried.

**Figure 1.**
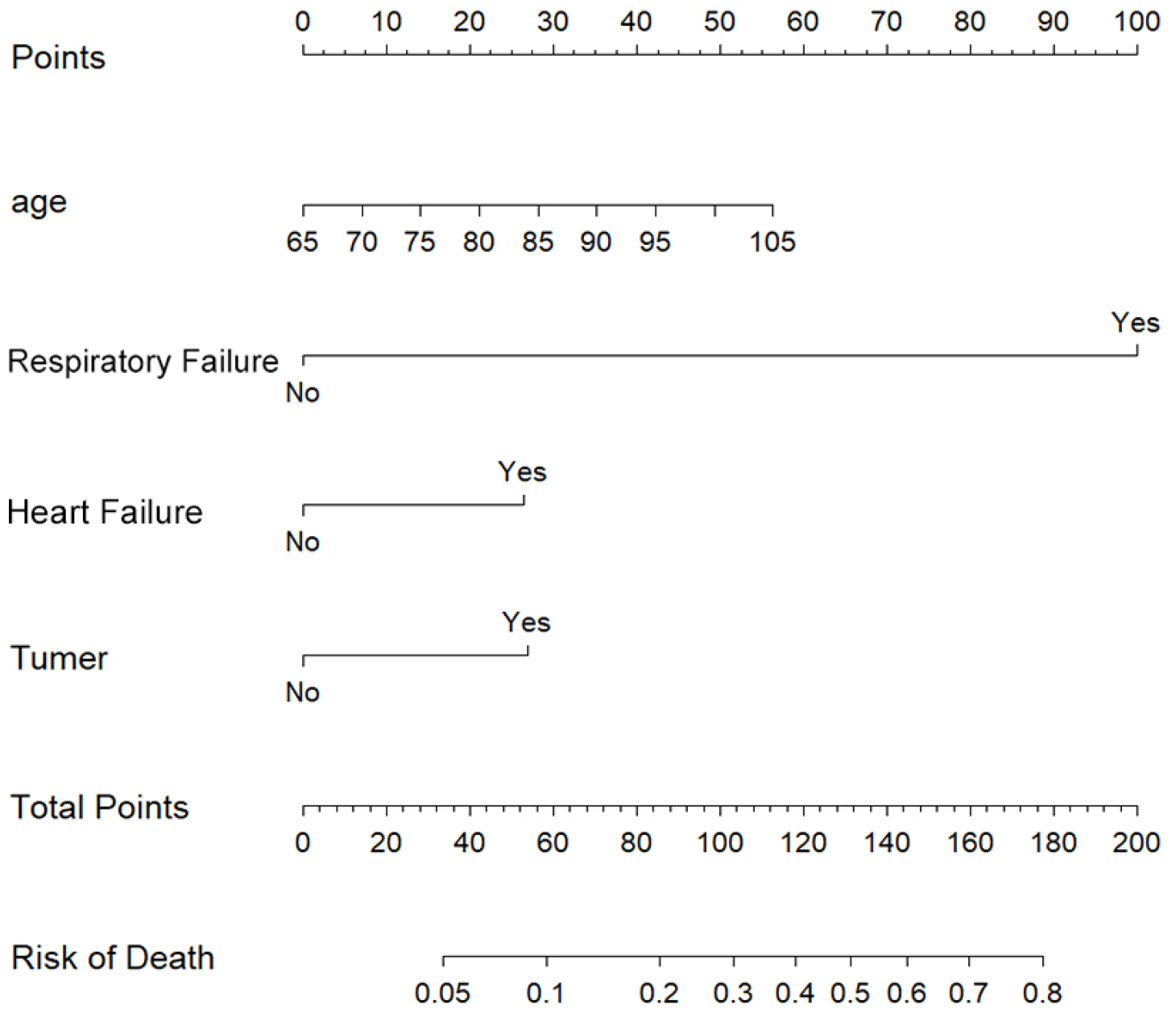
Nomogram based on the results of model to predict in-hospital mortality in elderly patients with CAP

### Model verification

According to the developed prediction model, the mortality rate of each patient in the training cohort was predicted and compared with the actual occurrence of in-hospital death [AUC = 0.877, 95% CI, 0.858–0.896; Figure 2(a)]. The calibration curve is shown in Figure 2(b). The calibration of the model satisfied the Hosmer–Lemeshow test (χ^2^ =5.681, *P* = 0.683). Thus, the calibration ability of the model is good.

**Figure 2.**
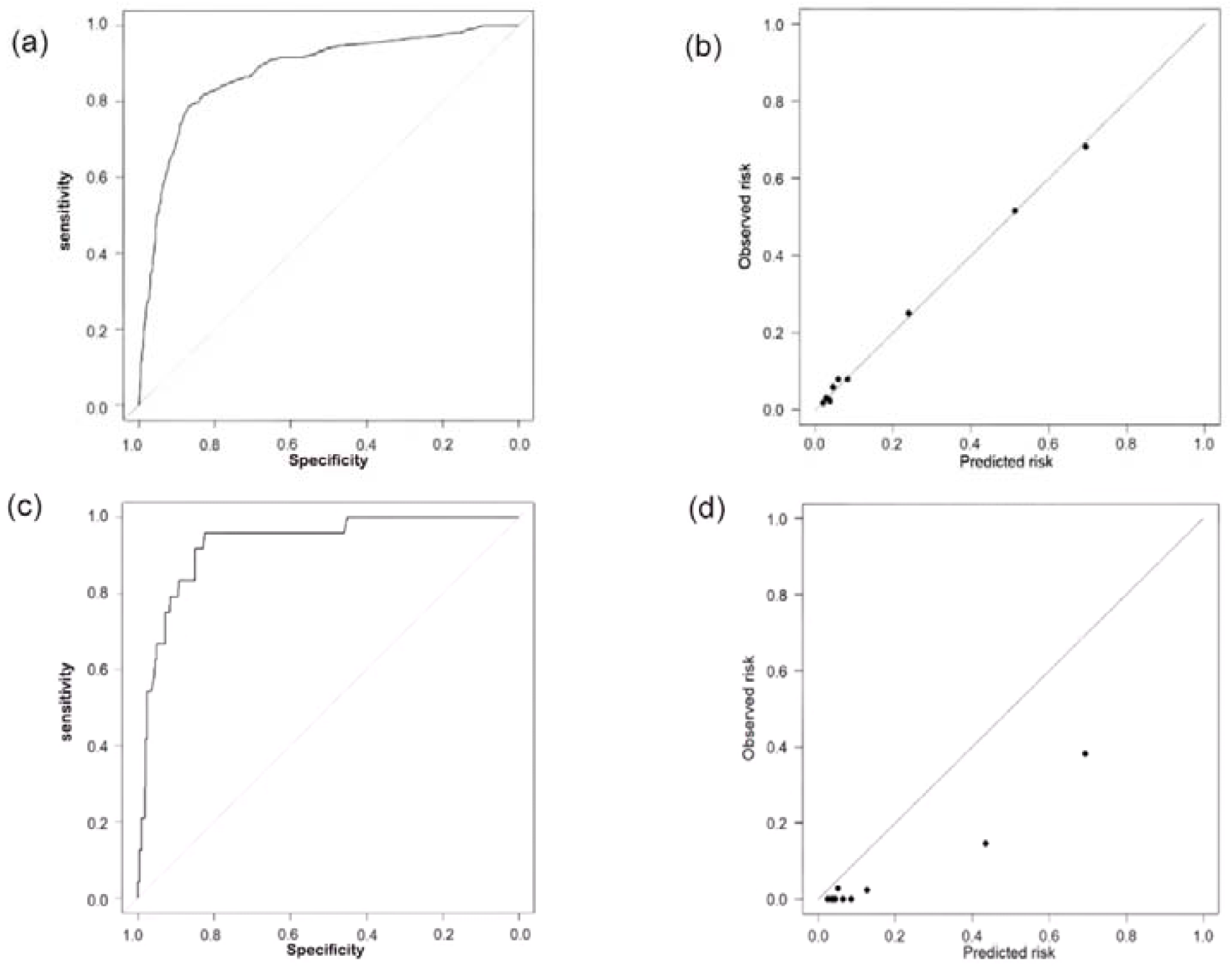
ROC curves of the training set (a) and validation set (c). Calibration curves of the training set (b) and validation set (d).

We utilized the validation set for external validation [Figures 2(c) and (d)]. The AUC of the validation set was 0.930, and the predictive ability of the model was high. However, the calibration curve showed that the actual probability (y-axis) was lower than the prediction probability (x-axis). This might result from the mild disease severity and low mortality rate in the validation set, leading to systematic error. By adjusting the cut-value, a better predictive accuracy can be achieved.

## DISCUSSION

Based on the data from the two cohorts, we used four predictors (age, respiratory failure, heart failure, and malignant tumors) to establish a model for predicting the risk of in-hospital death in elderly patients with CAP. The model was validated in a sizable validation cohort and achieved high accuracy. External Validation was conducted in a cohort from another hospital.

In this retrospective study of 2,890 elderly patients diagnosed with CAP in Beijing, we found an overall in-hospital mortality rate of 15.6%. In Sichuan, China, the in-hospital mortality rate of CAP patients over 65 years old was 10.1%.^6^

In this study, we developed a clinical predictive model to assess the prognosis in elderly patients with CAP in Beijing. Using logistic Cox regression, we identified four predictors (age, respiratory failure, heart failure, and malignant tumors) associated with poor prognosis in elderly patients with CAP. A clinically predictive model for assessing mortality was then developed based on the above four predictors.

Age is a well-known risk factor for CAP, especially in older people. Advanced age is associated with a higher risk of long-term mortality in CAP.^7, 8^ Calvillo-King et al. reviewed 20 studies on readmission and mortality after pneumonia.^9^ Most of these studies found that older age was associated with worse outcomes (age ≥ 65, OR = 2.7 [0.3–21.6]). Age may also interact with other factors in elderly people, including the presence of comorbidities, malnutrition, and cognitive impairment, all of which are related to the prognosis of elderly patients with CAP.^10^

One systematic review and meta-analysis of hospitalized patients with all-cause CAP found that the incidence rates for overall cardiac events, incident heart failure, acute coronary syndromes, and incident cardiac arrhythmias were 17.7%, 14.1%, 5.3%, and 4.7%, respectively;^11^ in the case of pneumococcal CAP, almost 20% of the studied patients had one or more of these cardiac events. These complications are associated with both short-term and long-term mortality.^12-14^

Acute respiratory failure and the development of sepsis are the main causes of CAP mortality.^15^ Long-term hypoxia caused by respiratory failure can lead to ischemic changes and dysfunction of the heart, brain, kidney, and lung and may worsen the outcome of CAP and/or lead to higher mortality.^16^

Almiral et al. found that the use of oral steroids and immunosuppressive therapy were clear risk factors for CAP, while the role of cancer was not clear.^17^ However, most patients with tumors use immunosuppressants and steroids as part of treatment. The increased risk of infectious complications is an important safety concern when prescribing immunosuppressive therapy. The prevention of infection is a key management strategy, and the early recognition of other associated risk factors for CAP should also be included as part of management.

As mentioned earlier, at least two clinical prediction tools (PSI and CURB-65) have been developed for stratifying patients with CAP based on risk.^4, 5^ Prospective comparisons of the predictive abilities of these two measures revealed c-statistics for predicting 30-day mortality ranging from 0.73–0.89 across studies.^18-20^ However, within any given study, no statistically significant differences were found between the two scales. The above two prediction models contain clinical information including test results, while some other models are based only on management data.

For example, Pine et al. reported a model based on ICD-9-CM codes with a c-statistic of 0.78.^21^ An administrative claims model developed to predict pneumonia mortality rates in hospitals based on patient age, sex, and 29 comorbidities (based on ICD-9-CM codes from the hospitalization and the prior year’s outpatient visits) produced a c-statistic of 0.72.^22^ In our study, the predictive model achieved a c-statistic of 0.877, significantly higher than those reported in previous studies. We also validated the model externally. The AUC of the validation set was 0.930, and the calibration curve indicated high predictive ability.

Our predictive model has several strengths. First, the clinical data collected were obtained from a large retrospective cohort over a 10-year period, resulting in good reliability. Second, the four predictors used in the model are routinely collected in clinical work; thus, the model can help healthcare workers determine the prognosis and outcomes of elderly patients of CAP, especially in resource-limited regions in China.

This study has several limitations. First, our main outcome was in-hospital mortality. However, the factors that are predictive of in-hospital mortality may differ from those that predict 30-day mortality, which has been used in other studies.^23^ It was impossible to study 30-day mortality in our study because the hospitalization time for some cases was less than 30 days. Second, all cases in this study were inpatient cases; outpatient and emergency patients were not included. Thus, our predictive model cannot be extended to elderly CAP patients in outpatient and emergency departments. Third, the model was only verified in two hospitals in Beijing. Further verification should be conducted in other areas and hospitals.

## CONCLUSION

In conclusion, we constructed a predictive model to evaluate mortality in elderly patients with CAP based on clinical data and information coded at discharge. This model provides a simple tool for physicians to assess the prognosis in elderly patients with CAP in Beijing.

## Data Availability

Date are available upon reasonable request to corresponding author.

## Acknowledgments

The authors thank AiMi Academic Services (www.aimieditor.com) for the English language editing and review services.

## Author contributors

YH, YY, YZ and FZ conceived and designed the study. YH and FZ collected data. HZ analysed and interpreted data. YH wrote the manuscript. All authors approved the final manuscript.

## Ethics approval

This study was carried out at the Peking University Third Hospital and Beijing Haidian Hospital. The study protocol was approved by the research ethics committee of Peking University Third hospital and complied with the principles of the Declaration of Helsinki(Project No. IRB00006761-M2021422). All data were used anonymously; written informed consent was not required because this was a retrospective study using deidentified clinical information.

## Provenance and peer review

Not commissioned; externally peer reviewed.

## Data availability statement

Date are available upon reasonable request to corresponding author.

